# A Prospective Study Measuring Resident/Faculty Contour Concordance: A Potential Tool for Quantitative Assessment of Residents’ Performance in Contouring and Target Delineation

**DOI:** 10.1101/2023.01.31.23285021

**Authors:** Caleb Nissen, Jun Ying, Faraz Kalantari, Mausam Patel, Arpan V. Prabhu, Anam Kesaria, Thomas Kim, Sanjay Maraboyina, Leslie Harrell, Fen Xia, Gary D. Lewis

## Abstract

**Purpose/Objective(s):** Accurate target delineation/contouring is essential for radiation treatment planning and radiotherapy efficacy. As a result, improving the quality of target delineation is an important goal in the education of radiation oncology residents. The purpose of this study was to track the concordance of radiation oncology residents’ contours with faculty physicians’ contours over the course of one year to assess for patterns.

**Materials/Methods:** Residents in PGY levels of 2-4 were asked to contour target volumes which were compared to the finalized, faculty physician-approved contours. Concordance between resident and faculty physician contours was determined by calculating the Jaccard Concordance Index (JCI), ranging from 0 or no agreement to 1 or complete agreement. Mutivariate mixed effect models were used to assess the association of JCI to the fixed effect of PGY level and its interactions with cancer type and other baseline characteristics. Post hoc means of JCI were compared between PGY levels after accounting for multiple comparisons using a Tukey’s method.

**Results:** In total, 958 structures from 314 patients collected during the 2020-2021 academic year were studied. The mean JCI was 0.77, 0.75, and 0.61 for the levels of PGY-4, PGY-3, and PGY-2, respectively. JCI score in PGY-2 was found lower than those of PGY-3 and PGY-4 respectively (P’s<0.001). No statistically significant difference of JCI score was found when comparing between PGY-3 and PGY-4 levels. The average JCI score was lowest (0.51) for primary head/neck cancers and highest (0.80) for gynecologic cancers.

**Conclusion:** Tracking and comparing the concordance of resident contours and faculty physician contours is an intriguing method of assessing resident performance in contouring and target delineation and could potentially serve as a quantitative metric in radiation oncology resident evaluation, which is lacking currently. However, additional study is necessary before this technique can be incorporated into residency assessments.

## Introduction

Accurate target delineation/contouring is essential for radiation treatment planning and the clinical efficacy of radiation therapy. Clinical trial data has demonstrated the importance of quality contouring and treatment planning on survival outcomes.^1^ As a result, improving the quality of contouring and target delineation is an important goal in the education of radiation oncology residents. There is limited quantitative data on the quality of residents’ contours, but previous studies indicate significant room for improvement.^2^ Therefore, it may be beneficial to track performance and improvement in resident contouring and target delineation during residency.

Advancements in radiation treatment planning software have allowed for easy collection and calculation of volumetric indices.^3^ Monitoring these indices during residency may serve as a quantifiable and objective way to track a resident’s contour quality during rotations. Currently, there are limited quantitative/objective measures to evaluate trainees’ clinical competency in radiation oncology residency. Current evaluations are based on clinical qualitative “Milestones” specified by the Accreditation Council for Graduate Medical Education (ACGME), which involve some degree of subjectivity and have been recently revised given critiques about variable interpretation by faculty and residents.^4–6^ We conducted a pilot study to track the concordance of radiation oncology residents’ contours with attending physicians’ contours to assess for patterns. We propose that concordance indices may be useful metrics to evaluate trainees’ performance in contouring and target delineation during radiation oncology residency.

## Materials and Methods

For this prospective study conducted during the 2020-2021 academic year, residents were asked to contour target volumes (gross target volume [GTV], clinical target volume [CTV], planning target volume [PTV]) based on patient history, physical exam, clinical stage and fused diagnostic imaging. They were allowed to use any available outside resources, including textbooks, review articles, consensus guidelines, and online atlases. Resident contours were saved as separate structures and compared to the finalized, board-certified attending physician-approved contours. All finalized contours and treatment plans went through standard departmental quality assurance protocols. Patients with all disease sites and treatment intents (curative or palliative) who underwent CT simulation from July 1, 2020, to June 30, 2021, were included. Patients with no saved resident/faculty contour structures were excluded. Contours from all residents (postgraduate year [PGY] levels 2-4) enrolled in our program were collected and analyzed. PGY-5 contours were not available for analysis as there was not a PGY-5 resident enrolled in our program during that year.

Contours were categorized by disease and patient characteristics. Cancers included in this study were head/neck, brain, genitourinary, gynecologic, thoracic, sarcoma, lymphoma, breast, and gastrointestinal. Data on patient gender, sex, cancer stage were collected from treatment notes in the electronic medical record. Additional information collected included whether the disease site being treated was the primary site versus metastatic site and if radiation was given definitively or adjuvantly (after surgery). All identifying patient information was anonymized. The Institutional Review Board (IRB) of the University of Arkansas for Medical Sciences (UAMS) gave ethical approval for this work.

Concordance between resident contours and attending physician contours was determined by calculating the Jaccard Concordance Index (JCI). The JCI was calculated by dividing the intersection volume of the resident/faculty contours by the union volume of the resident/faculty contours. A JCI score of 1 indicates complete agreement while a score of 0 indicates no agreement. While other concordance indices (such as the Dice Similarity Coefficient [DSC]) do exist, the JCI was chosen because it was easier to calculate given the tools available within Eclipse. Specifically, the Boolean Operations Tool in Eclipse (Varian Medical Systems, Palo Alto, CA, USA) was used to create the intersection volume of the resident/faculty contours as well as the union volume.

### Statistical analysis

The JCI score was considered as the primary outcome measure in this study, and it was defined as a numerical dependent variable. The key independent variable or fixed effect was the PGY level, a three-level categorical variable (PGY-2, PGY-3, and PGY-4) in this study. Other independent variables or fixed effects were cancer type, contour type, and treatment type. A multivariate mixed effect model was used to assess the association of JCI to the fixed effect of PGY, and its interaction with cancer type, after adjusting for baseline characteristics listed in Table 1. A random effect was used in the model to account for within-case correlation caused by multiple lesions in the same patient or case. Post hoc means were compared between PGY levels or groups under the mixed effect model framework and accounted for multiple comparisons using a Tukey’s method. Similar mixed effect models were used when PGY level interacted with another fixed effect such as contour type or treatment type. In order to further assess the association of JCI and PGY under interactions of both the cancer type and the contour type (or treatment type), a three-way mixed effect model was used after adjusting for the same baseline characteristics and accounting for within-case correlation using the same random effect. Again, post hoc means were compared between PGY levels in each sub-categorical group defined by the interaction of cancer type and contour type (or treatment type) after adjusting for multiple comparisons. Baseline categorical variables or characteristics were summarized using frequency (%). All statistical analyses were performed using SAS 9.4 software (SAS, Cary, NC). P-values <0.05 were considered statistically significant.

**Table 1:** Summary of Baseline Characteristics (N=314 patients)

| Variable | Group | Frequency (%) |
| --- | --- | --- |
| **Sex** | Female | 136 (43.31%) |
|  | Male | 178 (56.69%) |
| **Race** | White | 217 (69.11%) |
|  | Black | 78 (24.84%) |
|  | Other | 5 (1.59%) |
|  | Unknown | 14 (4.46%) |
| **Age** | ≤ 65 | 166 (53.21%) |
|  | > 65 | 146 (46.79%) |
| **Cancer Type/Site** | Primary Brain Tumors | 42 (13.38%) |
|  | Brain Metastases | 66 (21.02%) |
|  | Primary Breast Cancers | 15 (4.78%) |
|  | Primary Gastrointestinal Cancers | 29 (9.24%) |
|  | Primary Genitourinary Cancers | 27 (8.60%) |
|  | Primary Gynecologic Cancers | 18 (5.73%) |
|  | Primary Head & Neck Cancers | 38 (12.10%) |
|  | Lymphomas | 9 (2.87%) |
|  | Sarcomas | 4 (1.27%) |
|  | Primary Thoracic Cancers | 66 (21.02%) |
| **Primary/Metastatic Site** | Metastatic | 68 (21.73%) |
|  | Primary | 245 (78.27%) |
| **Stage** | I | 50 (23.47%) |
|  | II | 20 (9.39%) |
|  | III | 44 (20.66%) |
|  | IV | 99 (46.48%) |
| **Treatment Type** | Adjuvant | 99 (31.63%) |
|  | Definitive | 214 (68.37%) |
| **PGY Level** | 2 | 99 (31.53%) |
|  | 3 | 106 (33.76%) |
|  | 4 | 109 (34.71%) |
| **Contour Type\*** | CTV | 312 (32.57%) |
|  | GTV | 360 (37.58%) |
|  | PTV | 286 (29.85%) |

\*Contour Type frequency was obtained from a total of 958 contours/structures
PGY=Postgraduate Year
CTV=Clinical Target Volume
GTV=Gross Target Volume
PTV=Planning Target Volume

## Results

In total, 958 structures from 314 patients were available for analysis. Baseline patient, disease, treatment, and structure/contour characteristics are summarized in Table 1. The majority of patients were male (56.7%), white (69.1%), and under the age of 65 (53.2%). A large proportion of radiation treatments were given to the primary site (78.3%) as definitive treatment (68.4%). Mean JCI values stratified by baseline characteristic are summarized in Table 2. The mean JCI was 0.77, 0.75, and 0.61 for the PGY-4 level, PGY-3 level, and PGY-2 level, respectively. PGY levels 3 and 4 both had significantly greater JCI scores when compared to PGY-2 (P’s<0.001). No statistically significant variation was observed when comparing PGY-3 and PGY-4 between themselves. The average JCI scores across all PGY levels ranged from 0.51-0.80 on disease site subset analysis, with the lowest JCI score seen in primary head/neck cancers and the highest seen in gynecologic cancers. Head/neck cancers were found to have a significantly lower mean JCI as compared to primary brain tumors, metastatic brain tumors, primary gastrointestinal cancers, primary genitourinary cancers, primary gynecologic cancers, thoracic cancers, and lymphomas. There was a significant difference in mean JCI for metastatic versus primary site treatment (0.77 and 0.70 respectively, P=0.02) and adjuvant versus definitive treatment (0.65 and 0.74 respectively, P=0.001). The mean JCI for CTV structures (0.67) was significantly lower compared to the GTV (0.76, P<0.001) and PTV (0.74, P=0.001) structures. No significant difference was found for mean JCI as a function of patient race, age, or cancer stage.

**Table 2:** Summary of JCI values Stratified by Baseline Characteristics

| Variable | Group | Mean ± SE | P-value |
| --- | --- | --- | --- |
| Sex | *Female* | $0.75 \pm 0.02$ | *Reference* |
| | Male | $0.69 \pm 0.02$ | **0.0424** |
| Race | *White* | $0.72 \pm 0.02$ | *Reference* |
| | Black | $0.71 \pm 0.03$ | 0.9974 |
| | Other | $0.73 \pm 0.10$ | 0.9993 |
| | Unknown | $0.77 \pm 0.06$ | 0.8588 |
| Age | $\leq 65$ | $0.72 \pm 0.02$ | *Reference* |
| | $> 65$ | $0.71 \pm 0.02$ | 0.7153 |
| Cancer Type/Site | Primary Brain Tumors | $0.72 \pm 0.03$ | **0.0010** |
| | Brain Metastases | $0.77 \pm 0.03$ | **<0.0001** |
| | Primary Breast Cancers | $0.66 \pm 0.06$ | **0.3468** |
| | Primary Gastrointestinal Cancers | $0.75 \pm 0.04$ | **0.0003** |
| | Primary Genitourinary Cancers | $0.74 \pm 0.04$ | **0.0013** |
| | Primary Gynecologic Cancers | $0.80 \pm 0.05$ | **0.0002** |
| | *Primary Head & Neck Cancers* | $0.51 \pm 0.04$ | *Reference* |
| | Lymphomas | $0.78 \pm 0.07$ | **0.0253** |
| | Sarcomas | $0.75 \pm 0.11$ | 0.5100 |
| | Primary Thoracic Cancers | $0.73 \pm 0.03$ | **<0.0001** |
| Primary/Metastatic Site | *Metastatic* | $0.77 \pm 0.03$ | *Reference* |
| | Primary | $0.70 \pm 0.01$ | **0.0229** |
| Stage | I | $0.74 \pm 0.03$ | 0.9685 |
| | II | $0.70 \pm 0.05$ | 0.9895 |
| | III | $0.65 \pm 0.04$ | 0.3660 |
| | *IV* | $0.72 \pm 0.02$ | *Reference* |
| Treatment Type | *Adjuvant* | $0.65 \pm 0.02$ | *Reference* |
| | Definitive | $0.74 \pm 0.02$ | **0.0010** |
| PGY | 2 | $0.61 \pm 0.02$ | *Reference* |
| | 3 | $0.75 \pm 0.02$ | **<0.0001** |
| | 4 | $0.77 \pm 0.02$ | **<0.0001** |
| Contour Type | *CTV* | $0.67 \pm 0.02$ | *Reference* |
| | GTV | $0.76 \pm 0.02$ | **<0.0001** |
| | PTV | $0.74 \pm 0.02$ | **0.0012** |

JCI=Jaccard Concordance Index
SE=Standard Error
PGY=Postgraduate Year
CTV=Clinical Target Volume
GTV=Gross Target Volume
PTV=Planning Target Volume

On subset analysis of disease type, PGY levels 3 and 4 both had significantly greater JCI scores when compared to PGY-2 for primary gastrointestinal cancers (P=0.0039 and P<0.001), primary gynecologic cancers (P<0.001 and P=N/A [no PGY-4 data was available]), primary head/neck cancers (P=0.016 and P=0.029), and primary thoracic cancers (P=0.001 and P<0.001, respectively). There was no significant difference in mean JCI between PGY levels for primary brain tumors, metastatic brain tumors, primary breast cancers, genitourinary cancers, sarcomas, and lymphomas. This data is summarized in Table 3. On subset analysis of primary versus metastatic disease, PGY levels 3 (P<0.001) and 4 (P<0.001) both had significantly greater JCI scores compared to PGY-2 when contouring a primary site. In addition, PGY levels 3 (P<0.001) and 4 (P<0.001) both had significantly greater JCI scores compared to PGY-2 for definitive radiation treatment. This data is summarized in Table 4.

**Table 3:** Comparisons of JCI Values among PGY Levels Stratified by Cancer Type/Site

| Cancer Type/Site | Mean ± SE | | | P-value | | |
| --- | --- | --- | --- | --- | --- | --- |
|  | PGY-2 | PGY-3 | PGY-4 | PGY-2 vs PGY-3 | PGY-2 vs PGY-4 | PGY-3 vs PGY-4 |
| Brain Metastases | 0.78 ± 0.04 | 0.70 ± 0.05 | 0.81 ± 0.04 | 0.4649 | 0.8953 | 0.2510 |
| Primary Breast Cancers | NA | 0.67 ± 0.08 | 0.65 ± 0.13 | NA | NA | 0.9025 |
| Primary Gastrointestinal Cancers | 0.53 ± 0.05 | 0.82 ± 0.07 | 0.89 ± 0.04 | **0.0039** | **<0.0001** | 0.6482 |
| Primary Genitourinary Cancers | 0.63 ± 0.05 | 0.80 ± 0.05 | 0.81 ± 0.05 | 0.0577 | **0.0383** | 0.9935 |
| Primary Gynecologic Cancers | 0.57 ± 0.06 | 0.89 ± 0.04 | NA | **0.0005** | NA | NA |
| Primary Head & Neck Cancers | 0.38 ± 0.05 | 0.57 ± 0.04 | 0.55 ± 0.04 | **0.0155** | **0.0294** | 0.9717 |
| Lymphomas | NA | 0.84 ± 0.21 | 0.78 ± 0.07 | NA | NA | 0.7860 |
| Primary Brain Tumors | 0.66 ± 0.06 | 0.76 ± 0.07 | 0.74 ± 0.07 | 0.5278 | 0.6697 | 0.9643 |
| Sarcomas | 0.26 ± 0.12* | 0.91 ± 0.09 | 0.92 ± 0.12* | 0.2125 | 0.2408 | 0.9984 |
| Primary Thoracic Cancers | 0.57 ± 0.05 | 0.81 ± 0.04 | 0.82 ± 0.05 | **0.0013** | **0.0008** | 0.9679 |

\*N=1 patient
JCI=Jaccard Concordance Index
PGY=Postgraduate Year
SE=Standard Error
NA=No Available contours/structures

**Table 4:**
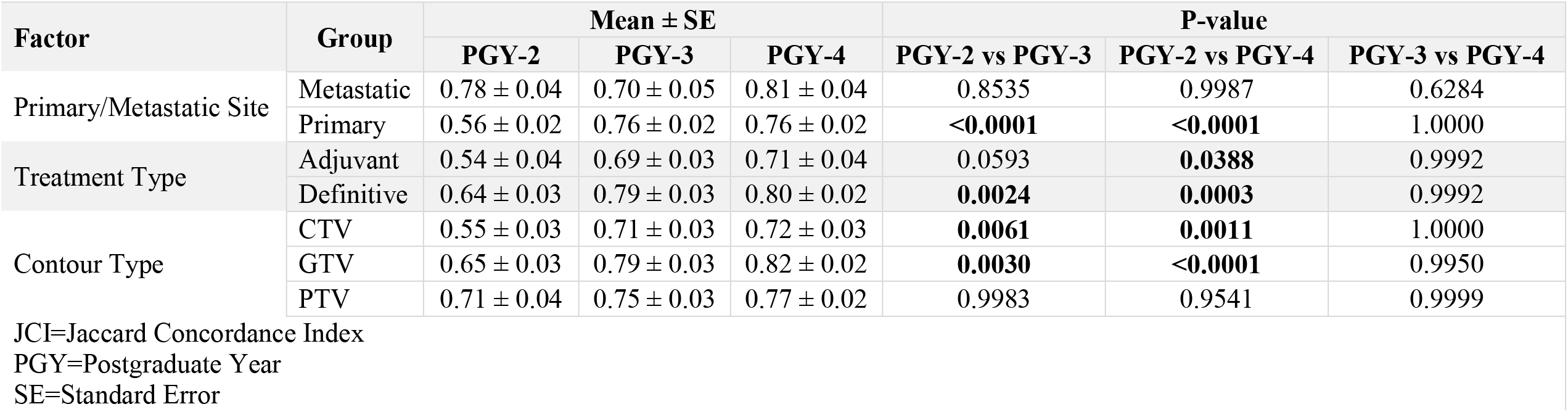
Comparisons of Means of JCI Values among PGY Levels Stratified by Treatment Characteristics

| Factor | Group | Mean ± SE |  |  | P-value |  |  |
| --- | --- | --- | --- | --- | --- | --- | --- |
|  |  | PGY-2 | PGY-3 | PGY-4 | PGY-2 vs PGY-3 | PGY-2 vs PGY-4 | PGY-3 vs PGY-4 |
| Primary/Metastatic Site | Metastatic | 0.78 ± 0.04 | 0.70 ± 0.05 | 0.81 ± 0.04 | 0.8535 | 0.9987 | 0.6284 |
|  | Primary | 0.56 ± 0.02 | 0.76 ± 0.02 | 0.76 ± 0.02 | **<0.0001** | **<0.0001** | 1.0000 |
| Treatment Type | Adjuvant | 0.54 ± 0.04 | 0.69 ± 0.03 | 0.71 ± 0.04 | 0.0593 | **0.0388** | 0.9992 |
|  | Definitive | 0.64 ± 0.03 | 0.79 ± 0.03 | 0.80 ± 0.02 | **0.0024** | **0.0003** | 0.9992 |
| Contour Type | CTV | 0.55 ± 0.03 | 0.71 ± 0.03 | 0.72 ± 0.03 | **0.0061** | **0.0011** | 1.0000 |
|  | GTV | 0.65 ± 0.03 | 0.79 ± 0.03 | 0.82 ± 0.02 | **0.0030** | **<0.0001** | 0.9950 |
|  | PTV | 0.71 ± 0.04 | 0.75 ± 0.03 | 0.77 ± 0.02 | 0.9983 | 0.9541 | 0.9999 |

JCI=Jaccard Concordance Index
PGY=Postgraduate Year
SE=Standard Error

## Discussion

This single institutional pilot study demonstrated a statistically significant difference in mean JCI scores between upper-level radiation oncology residents (PGY levels 3 and 4) and junior PGY-2 residents when evaluating target structures across all disease sites. This observed difference in JCI values likely reflects the increased experience with contouring that upper-level residents have compared to junior residents. As such, the quality of resident contouring and target delineation over the course of residency training may potentially be evaluated quantitatively by using JCI scores (or other similar concordance indices).

Currently, there are a lack of quantitative and objective metrics to assess resident competence during radiation oncology residency. While this is not unique to the field of radiation oncology,^7,8^ the fact remains that it can be difficult to standardize resident assessments. This lack of objective/quantitative assessment tools also makes it challenging to interpret and compare competency standards across programs. Right now, the bulk of resident assessments is centered on the clinical “Milestones” specified by the ACGME, which involve some degree of subjectivity^9,10^ and have been recently revised given critiques about complex language, lack of clear progression between PGY levels, and variable interpretation by both faculty and residents.^4–6^ One objective measure currently used is the number of CT simulations participated in or observed by a resident. However, a case may be logged by a trainee but does not necessarily specify the degree of resident involvement or the quality of resident performance.

Reflecting this desire from stakeholders for improved radiation oncology residency program training and assessment, the ACGME and the Review Committee for Radiation Oncology (RORC) recently increased standards for residency programs by increasing the number of required minimum cases by disease site and increasing the time spent by residents at the main educational site.^11^ We propose that tracking resident/faculty contour concordance provides another objective tool to measure, follow, and assess resident performance during residency and can be used as complementary information when combined with the ACGME Milestones and case log numbers.

Tracking resident/faculty concordance may also allow for targeted initiatives to improve contouring and target delineation for specific residents, disease sites, or treatment types. For example, our study found variation in mean JCI scores among all residents for different anatomical subsites. The lowest JCI score was 0.51 for primary head/neck cancers. Primary breast cancers also had a relatively low JCI of 0.66. The highest JCI scores were found for primary gynecologic cancers, primary genitourinary cancers, metastatic brain cancer, and lymphomas. For these sites, the mean JCI scores ranged from 0.77 to 0.82. This data was somewhat expected as head/neck cancers are considered to be the most difficult to contour, while brain metastases are relatively straightforward to distinguish on planning MRI. This information could be used to implement additional training initiatives (such as head/neck contouring lectures or practice workshops) and measure the effect on JCI scores during residency. JCI scores also tended to be lower in other complex contouring situations. For example, the mean JCI score when treating the primary site was statistically significantly lower (possibly due to the need to contour both the tumor and draining lymphatics) compared to treating a metastatic site which can be relatively uncomplicated (as is the case when contouring brain metastases). Similarly, the mean JCI score for adjuvant radiation was statistically significantly lower (possibly due to the difficulty in delineating anatomy after surgery) compared to definitive radiation.

There has been very limited research performed evaluating the utility of the JCI (or other concordance indices) as a quantitative measures of resident contour quality over the course of residency training. There have been several studies using concordances indices such as the JCI, DSC, overlap volume (OV), volume difference, mean surface distance (MSD), and simultaneous truth and performance level estimation (STAPLE) to analyze resident contour quality before and after an educational intervention (course, workshop, contour atlas, computer atlas, etc.),^12–18^ indicating the value of these educational tools. To our knowledge, our study is the first to serially monitor and measure resident contour quality during residency training. The large number of target structures (>900) analyzed is another strength of our study. The results of this pilot study may provide the basis for a larger prospective study in partnership with other institutions or the ACGME/RORC.

There are several limitations to our study. First, the results were obtained from a single radiation oncology department and may not be representative of other departments. In addition, attending contours were chosen to represent the “gold standard,” given each attending’s board-certification and multiple department quality assurance protocols. However, there is likely to be variability among attending physician contours, as has been demonstrated in multi-institutional clinical trial quality assurance protocols.^19^ These limitations highlight the need for larger prospective studies across multiple institutions.

## Conclusion

In this single-institutional prospective study, we demonstrated that measuring and monitoring resident contouring and target delineation performance during residency training using the Jaccard Concordance Index was feasible. Tracking and comparing the concordance of resident contours and faculty physician contours could potentially serve as a quantitative objective metric in radiation oncology resident evaluation, which is lacking currently. However, additional study is necessary before this technique can be incorporated into residency assessments.

## Data Availability

Research data are not available at this time.

## [Acknowledgements]

The study was supported by the Biostatistics Shared Resource of University of Arkansas for Medical Sciences Winthrop P. Rockefeller Cancer Institute.

## Notes

**[Conflict of Interest Statement for All Authors]** The authors have no conflicts of interest to disclose.

### Competing Interest Statement

The authors have declared no competing interest.

### Funding Statement

This study did not receive any funding

### Author Declarations

The Institutional Review Board (IRB) of the University of Arkansas for Medical Sciences (UAMS) gave ethical approval for this work.

